# Changes in HIV Treatment Differentiated Care Utilization During the COVID-19 Pandemic in Zambia

**DOI:** 10.1101/2021.03.20.21254021

**Authors:** Youngji Jo, Sydney Rosen, Karla Therese L Sy, Bevis Phiri, Amy N Huber, Muya Mwansa, Hilda Shakwelele, Prudence Haimbe, Mpande M Mwenechanya, Priscilla Lumano Mulenga, Brooke E Nichols

## Abstract

**Background:** Differentiated service delivery (DSD) models aim to lessen the burden of HIV treatment on patients and providers in part by reducing requirements for facility visits and extending dispensing intervals. With the advent of the COVID-19 pandemic, minimizing patient contact with healthcare facilities and other patients, while maintaining treatment continuity and avoiding loss to care, has become more urgent, resulting in efforts to increase DSD uptake. We assessed the extent to which DSD coverage and antiretroviral treatment (ART) dispensing intervals have changed during the COVID-19 pandemic in Zambia.

**Methods:** We used patient data from Zambia’s electronic medical record system (SmartCare) for 737 health facilities, representing about 3/4 of all ART patients nationally, to compare the numbers and proportional distributions of patients enrolled in DSD models in the six months before and six months after the first case of COVID-19 was diagnosed in Zambia in March 2020. Segmented linear regression was used to determine whether the introduction of COVID-19 into Zambia further accelerated the increase in DSD scale-up.

**Results:** Between September 2019 and August 2020, 181,317 patients aged 15+ (81,520 and 99,797 from September 1, 2019 to March 1, 2020 and from March 1 to August 31, 2020, respectively) enrolled in DSD models in Zambia. Overall participation in all DSD models increased over the study period, but uptake varied by model. The rate of acceleration increased in the second period for home ART delivery (152%), ≤2-month fast-track (143%), and 3-month MMD (139%). There were significant decelerations in the increase in enrolment for 4-6-month fast-track (−28%) and ‘other’ models (−19%).

**Conclusions:** Participation in DSD models for stable ART patients in Zambia increased after the advent of COVID-19, but dispensing intervals diminished. Eliminating obstacles to longer dispensing intervals, including those related to supply chain management, should be prioritized to achieve the expected benefits of DSD models and minimize COVID-19 risk.

## Introduction

In 2020, an estimated 16.4 million people taking antiretroviral therapy (ART) for HIV treatment in sub-Saharan Africa risked treatment interruptions because of COVID-19 due to closing or limiting of HIV services, antiretroviral supply chain disruptions, and/or overwhelmed service providers.[1] Maintenance of ART services — in addition to continued case identification and prompt initiation of newly diagnosed people living with HIV (PLHIV) on lifelong treatment — is critical to protect the progress that has been made towards HIV epidemic control.[2]

One potential solution to the disruptions caused by COVID-19 is differentiated service delivery (DSD), a “patient-centered approach that simplifies and adapts HIV services across the cascade to serve the needs of people living with HIV (PLHIV) better and reduce unnecessary burdens on the health system.”[3] DSD has emerged as a key strategy of HIV programs in resource-limited settings, as, DSD models can lessen the burden of HIV treatment on patients and providers in part by extending medication dispensing intervals, reducing requirements for facility visits, and adjusting the location of service delivery. These adjustments also minimize patient contact with healthcare facilities and other patients, a high priority during the COVID-19 pandemic.

In Zambia, the Ministry of Health began promoting DSD models for antiretroviral treatment in 2016, with participation gradually increasing over time.[4] By February 2021, roughly a quarter of the country’s nearly 1 million ART patients had ever been enrolled in a DSD model. The models offered in Zambia included multi-month dispensing (MMD), fast-track medication pickup, community adherence groups (CAGs), and home ART delivery, with healthcare facilities varying widely on which of these or other models they adopted. Since 2016, 3-month dispensing has been the standard of care for stable patients, though it has not been universally implemented. Six-month dispensing was introduced in 2019. When the country’s first SARS-CoV-2 infection was confirmed in March 2020, the Ministry of Health doubled down on implementation of 3- and 6-month dispensing for stable patients. Other models became more or less attractive in the face of COVID-19 risks and restrictions, depending on whether they required patients to meet as groups (e.g. CAGs) or reduced the need for public interaction (e.g. home delivery). In this study, we assessed the association between the COVID-19 pandemic and Zambia’s response to it and the rate of change of enrolment in DSD models in the six-month period before and after diagnosis of the first SARS-CoV-2 case.

## Methods

To assess how DSD model enrolment, by model type, changed before and after the start of the COVID-19 pandemic, we conducted a retrospective review of SmartCare, Zambia’s national electronic medical record system. As of February 2021, 737,411 patients were recorded in SmartCare as currently on ART, representing roughly three quarters of all ART patients in the country. The remaining quarter of patients attend facilities that do not yet utilize SmartCare. We accessed records for all patients aged 15 years or older who enrolled in any DSD model between September 2019 and August 2020 at any of 737 health facilities across all 10 provinces. We collapsed the many DSD models recorded in SmartCare into eight groups based on the location and duration of medication dispensing: ≤2-month fast-track, 3-month fast-track, 4-6-month fast-track, 3-month MMD, 4-6-month MMD, CAGs, home ART delivery, and all others. A description of each model can be found in Table 1).[5]

**Table 1.**
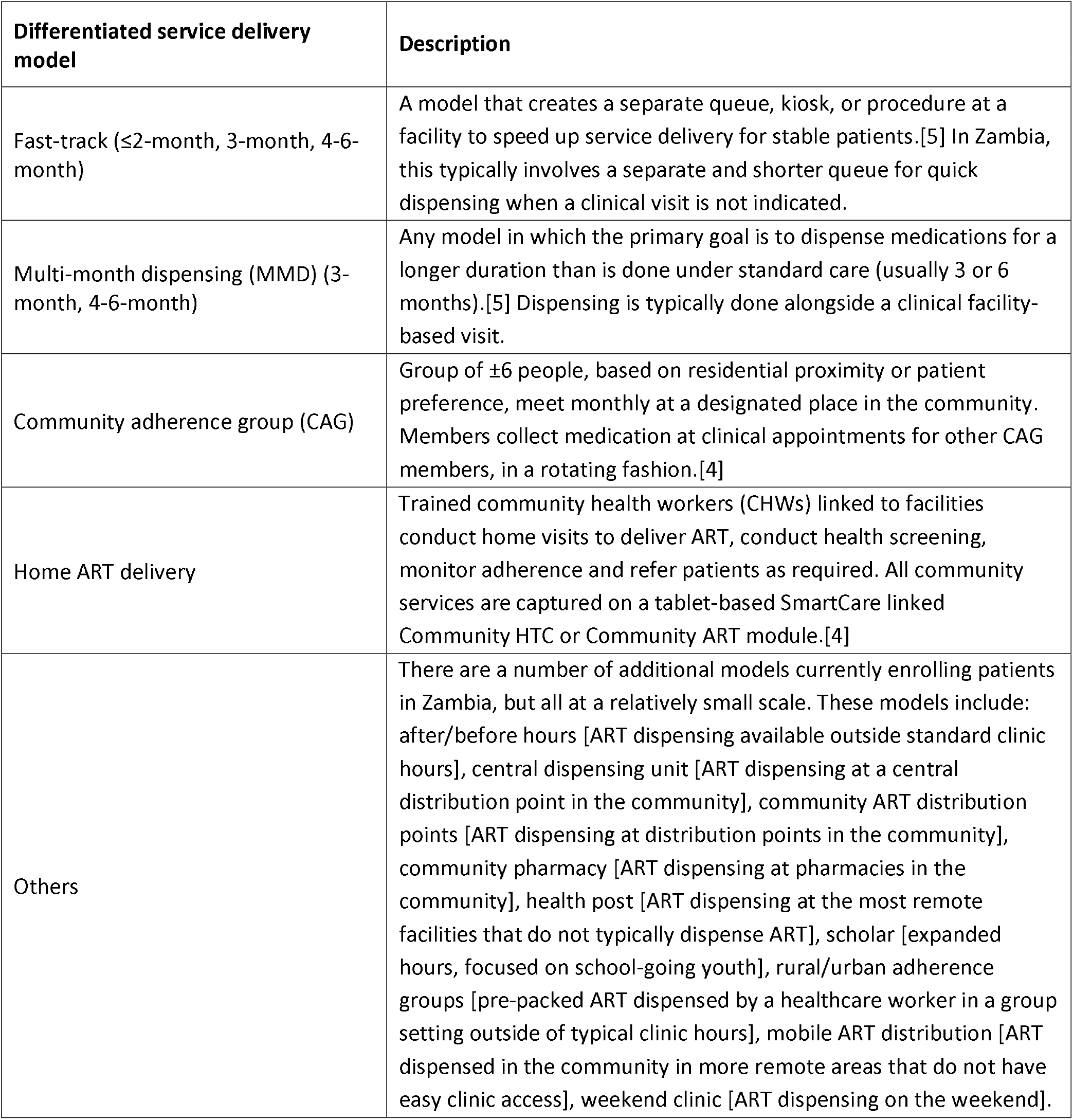
Description of each of the evaluated differentiated service delivery models implemented in Zambia between September 2019 and August 2020

| Differentiated service delivery model | Description |
| --- | --- |
| Fast-track (≤2-month, 3-month, 4-6-month) | A model that creates a separate queue, kiosk, or procedure at a facility to speed up service delivery for stable patients.[5] In Zambia, this typically involves a separate and shorter queue for quick dispensing when a clinical visit is not indicated. |
| Multi-month dispensing (MMD) (3-month, 4-6-month) | Any model in which the primary goal is to dispense medications for a longer duration than is done under standard care (usually 3 or 6 months).[5] Dispensing is typically done alongside a clinical facility-based visit. |
| Community adherence group (CAG) | Group of ±6 people, based on residential proximity or patient preference, meet monthly at a designated place in the community. Members collect medication at clinical appointments for other CAG members, in a rotating fashion.[4] |
| Home ART delivery | Trained community health workers (CHWs) linked to facilities conduct home visits to deliver ART, conduct health screening, monitor adherence and refer patients as required. All community services are captured on a tablet-based SmartCare linked Community HTC or Community ART module.[4] |
| Others | There are a number of additional models currently enrolling patients in Zambia, but all at a relatively small scale. These models include: after/before hours [ART dispensing available outside standard clinic hours], central dispensing unit [ART dispensing at a central distribution point in the community], community ART distribution points [ART dispensing at distribution points in the community], community pharmacy [ART dispensing at pharmacies in the community], health post [ART dispensing at the most remote facilities that do not typically dispense ART], scholar [expanded hours, focused on school-going youth], rural/urban adherence groups [pre-packed ART dispensed by a healthcare worker in a group setting outside of typical clinic hours], mobile ART distribution [ART dispensed in the community in more remote areas that do not have easy clinic access], weekend clinic [ART dispensing on the weekend]. |

We first describe the basic characteristics of patients enrolled by DSD model before and after the introduction of COVID-19 in Zambia to determine whether enrolment in models has changed in terms of location (urban/rural, level of health facility) or in the age or sex distribution of patients enrolling. For each of the DSD model groups, we calculated the number of DSD enrolments by month from September 2019 to August 2020. To assess the effect of the COVID-19 pandemic on DSD care utilization, we conducted an interrupted time series analysis using a segmented regression. Segmented regression has been previously used to evaluate changes at any defined point in time.[6] In our analysis, we compared the change in slope between the cumulative number of patients enrolled in DSD before March 1, 2020 compared to March 1^st^ through August 2020 (i.e. before and after March 1st, 2020), the approximate date when COVID-19 was first diagnosed in Zambia.[7] We used the following segmented regression model: *DSD* = *β*_0_ + *β*_1_ *time + β*_*2*_*covid*_*t*_ *+β*_*3*_*time*.*covid*_*t*_ where time is in months, and covid is a dummy variable indicating whether the current time is pre- or post-covid. The outcome *DSD* is the cumulative number of patients enrolled in DSD at time t. *β*_*3*_ indicates the slope change following the intervention, which we then tested whether there was a significant change in *β*_*3*_ before and after March 1, 2020; a significant change in slope would suggest that DSD utilization changed substantially during the COVID-19 pandemic. All analyses were performed at a two-sided significance level of 0.05. Finally, we estimated percent changes in participation between the periods for each model group based on the mean slope. Data analysis was conducted in *R version 4*.*0*.*2*. (The R Project for Statistical Computing, Vienna Austria).

## Ethics

This study protocol was approved by ERES Converge IRB (Zambia), protocol number 2019-Sep-030, the Human Research Ethics Committee (Medical) of the University of Witwatersrand, protocol number M190453, and the Boston University IRB H-38823.

## Results

Participation in DSD models before and after the introduction of COVID-19 in March 2020 is presented in Table 2. Between September 2019 and August 2020, 181,317 patients aged 15+ enrolled in DSD models in Zambia, including 81,520 before and 99,797 on or after March 1, 2020, an overall increase of 22.4%. Uptake varied widely by model, however. Between the two periods, participation in 3-month dispensing increased from 13% to 21% of all DSD enrollments, ≤2-month fast-track from 7% to 10%, and home ART delivery from 1% to 2%. The proportion of all DSD enrollments in 4-6-month fast-track fell from 23% to 14%, 3-month fast-track from 8% to 7%, and CAGs from 4% to 3%. There was no change in the proportion enrolled in 4-6-month MMD (38% of all DSD enrolments in both periods). Proportions of patients enrolled in rural areas increased for ≤3-month fast-track, 3-month-MMD, community adherence groups, and others. Home ART delivery was the only model to see a relative increase in the proportion of patients enrolled in urban areas (Table 2). There were no significant differences between the two time periods in the composition of the population enrolled in terms of sex or age.

**Table 2.** Percentage change in numbers of patients enrolled in DSD models before and after COVID-19 introduction in Zambia (n=181,317)

| Parameters |  | Number of patients | Location |  | Health care level |  |  | Sex |  | Age (Mean, SD) |
| --- | --- | --- | --- | --- | --- | --- | --- | --- | --- | --- |
|  |  |  | Urban | Rural | Health post | Clinic | Hospital | Male | Female |  |
| All models | Before<sup>a</sup> | 81520 | 64997 | 16523 | 4587 | 49619 | 27314 | 28562 | 46808 | 42 (11) |
|  | After<sup>b</sup> | 99797 | 75424 | 24373 | 5218 | 58403 | 36176 | 36386 | 59587 | 41 (12) |
|  | %Δ<sup>c</sup> | 22% | 16% | 48% | 14% | 18% | 32% | 27% | 27% | - |
| ≤2-month fast track dispensing | Before | 6005 | 4405 | 1600 | 302 | 4202 | 1501 | 2078 | 3927 | 40 (12) |
|  | After | 10163 | 7205 | 2958 | 618 | 6854 | 2691 | 3654 | 6509 | 39 (11) |
|  | %Δ | 69% | 64% | 85% | 105% | 63% | 79% | 76% | 66% | - |
| 3-month fast-track dispensing | Before | 6325 | 5788 | 537 | 318 | 3973 | 2034 | 2067 | 4258 | 41 (10) |
|  | After | 6917 | 6140 | 777 | 350 | 4360 | 2207 | 2427 | 4490 | 41 (11) |
|  | %Δ | 9% | 6% | 45% | 10% | 10% | 9% | 17% | 5% | - |
| 4-6-month fast-track dispensing | Before | 19112 | 18283 | 829 | 2013 | 10026 | 7073 | 6481 | 12631 | 43 (10) |
|  | After | 14168 | 13627 | 541 | 975 | 7553 | 5640 | 5172 | 8996 | 43 (10) |
|  | %Δ | -26% | -25% | -35% | -52% | -25% | -20% | -20% | -29% | - |
| 3-month MMD | Before | 10743 | 8215 | 2528 | 410 | 7101 | 3232 | 3744 | 6999 | 41 (11) |
|  | After | 21101 | 14812 | 6289 | 1122 | 13030 | 6949 | 7564 | 13537 | 41 (12) |
|  | %Δ | 96% | 80% | 149% | 174% | 83% | 115% | 102% | 93% | - |
| 4-6-month MMD | Before | 30832 | 22447 | 8385 | 998 | 19689 | 10145 | 11246 | 19586 | 44 (11) |
|  | After | 38120 | 28260 | 9860 | 1439 | 21576 | 15105 | 14172 | 23948 | 43 (11) |
|  | %Δ | 24% | 26% | 18% | 44% | 10% | 49% | 26% | 22% | - |
| Community adherence groups | Before | 2885 | 1595 | 1290 | 112 | 1628 | 1145 | 917 | 1968 | 45 (11) |
|  | After | 3483 | 1362 | 2121 | 133 | 2220 | 1130 | 1231 | 2252 | 45 (11) |
|  | %Δ | 21% | -15% | 64% | 19% | 36% | -1% | 34% | 14% | - |
| Home ART delivery | Before | 721 | 444 | 277 | 240 | 132 | 349 | 283 | 438 | 39 (12) |
|  | After | 1929 | 1472 | 457 | 288 | 838 | 803 | 686 | 1243 | 39 (12) |
|  | %Δ | 168% | 232% | 65% | 20% | 535% | 130% | 142% | 184% | - |
| Others | Before | 4897 | 3820 | 1077 | 194 | 2868 | 1835 | 1746 | 3151 | 40 (13) |
|  | After | 3916 | 2546 | 1370 | 293 | 1972 | 1651 | 1480 | 2436 | 39 (13) |
|  | %Δ | -20% | -33% | 27% | 51% | -31% | -10% | -15% | -23% | - |

a. Before- September 2019-February 2020
b. After- March 2020-August 2020
c. Percentage increase between before and after periods

Participation in DSD models accelerated over the study period. Comparing the periods before and after March 1, 2020, segmented linear regression models demonstrated an acceleration in the rate of increase (significant increases in slope) in participation during the COVID-19 pandemic for home ART delivery (152% change in slope between periods), ≤2-month fast-track (143%), and 3-month MMD (139%). Three-month fast-track showed both an immediate increase in numbers enrolled (155% from 6,278 to 9,729) and a significant acceleration in the rate of increase (60%) between the two periods. In contrast, there were significant decelerations in the increase in enrolment for 4-6-month fast-track (−28%) and for ‘other’ models (−19%) (Figure 1).

**Figure 1.**
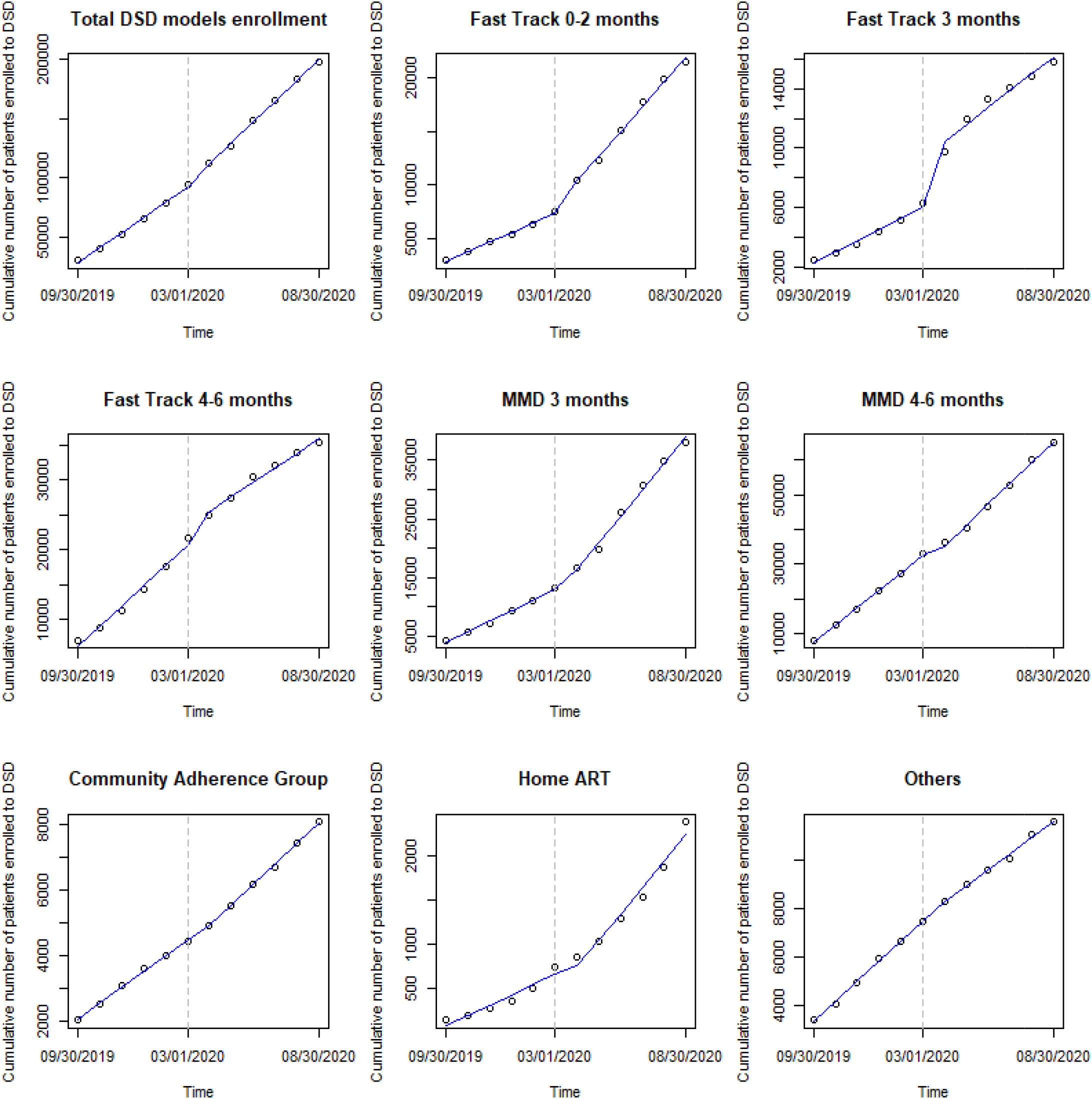
Interrupted time series scatter plot and slope lines for the DSD models before (September 2019-February 2020) and after (March 2020-August 2020) March 1, 2020 in Zambia

## Discussion

Over the course of 2020, the COVID-19 pandemic was associated with accelerated participation in DSD models in Zambia, though with uneven increases across the models. Most new patients enrolled in ≤2-month fast track, 3-month MMD, 4-6-month MMD, CAGs, or home ART delivery. On the other hand, increase in DSD enrollment was slower for the 4-6-month fast-track and ‘other’ models. Participation in home ART delivery increased the most (168%), but it still accounted for only a small proportion of all participation (2%). Recommendations that high-risk individuals remain at home, in order to minimize their exposure to SARS-CoV-2, may potentially explain the expansion of home delivery models. We also found an immediate jump in enrolment for ≤2-month and 3-month fast-track in March 1, 2020 and an increase one month later for home ART delivery.

Although 6-month dispensing is Zambia’s national policy for stable patients, the proportion of patients enrolled in the 6-month dispensing model fell between the two time periods, while ≤3-month dispensing increased. The shortening of dispensing intervals may be primarily due to patients switching temporarily from the newer ARV regimen of tenofovir+lamivudine+dolutegravir (TLD) back to the previous regimen tenofovir+lamivudine+efavirenz (TLE) to mitigate threats to the TLD global supply chain. In a recent WHO survey, five of 13 countries in sub-Saharan Africa reported ART stock availability for major first-line regimens to be 3 months or less due, in part, to failure of suppliers to deliver on time.[8] There is clear tension between the desire to implement longer-duration dispensing to minimize patient interaction at healthcare facilities due to COVID-19 transmission concerns and the reality of the impact that the pandemic has had on the global supply chain. If supply chain concerns are responsible for the drop in dispensing duration we observed in Zambia, then 6-month dispensing should be expected to rebound as the pandemic is brought under control and the supply chain strengthens.

Our study has several limitations. We relied entirely on routinely collected medical record data, which may be incomplete in some cases. While interrupted time series analysis allows the ability to control for secular trends in the data (unlike pre-/post-cross-sectional studies) using population-level data with clear graphical presentation of results, this analysis does not illustrate how and why the introduction of COVID-19 resulted different scale-up patterns by DSD models and whether and to what extent the temporal changes may differ by setting. Future research may examine the drivers and barriers of multi-month dispensing from both the demand and supply side aspects in the context of COVID-19 to improve continuation of care. Moreover, this analysis focused on DSD enrolment only; we have not considered retention in the DSD models or care more generally. Future work should aim to understand how this rapid acceleration of DSD uptake has affected overall retention in care.

## Conclusions

Based on national electronic medical record data for patients enrolled in DSD models in Zambia from September 2019-August 2020, our findings suggest that the introduction of the COVID-19 pandemic was associated with an acceleration in the scale-up of DSD models for patients on ART in Zambia. Efforts to eliminate obstacles to longer dispensing intervals should be prioritized to achieve the expected benefits of DSD models and minimize COVID-19 risk. This process has already begun in Zambia, where the government is now recommending relaxation of eligibility criteria for multi-month dispensing, such that all patients initiating ART to receive a three or six month supply of medications immediately, allowing them to delay their first follow-up visit for three or six months after initiation [9]. Evaluating the impact of this evolution in DSD guidelines will be a high priority for the coming years.

## Data Availability

Raw data were obtained from Zambia's national electronic medical record system. Derived data supporting the findings of this study are available from the corresponding author [BEN] on request.

## Competing interests

The authors declare that they have no conflicting interests.

## Authors’ contributions

YJ, SR and BEN conceived the study. YJ, SR, KTLS, BEN designed the study. BP, MM, HS, PH, MMM, PLM led study data collection. YJ, KTLS analyzed the data, and SR, ANH, BEN contributed to data analysis. YJ, SR, KTLS, BEN wrote the first draft of the manuscript. All authors reviewed and edited the manuscript. All authors have read and approved the final manuscript.

## Acknowledgements

None declared

## Funding

Funding for the study was provided by the Bill & Melinda Gates Foundation through OPP1192640 to Boston University. The funder had no role in study design, data collection and analysis, decision to publish or preparation of the manuscript.

## List of abbreviations

ART: Antiretroviral therapy
CAG: Community adherence group
DSD: Differentiated service delivery
HIV: Human immunodeficiency virus
MMD: Multi-month dispensing

## Notes

### Competing Interest Statement

The authors have declared no competing interest.

